# Clinical Metagenomic Sequencing for Species Identification and Antimicrobial Resistance Prediction in Orthopaedic Device Infection

**DOI:** 10.1101/2021.08.13.21261967

**Authors:** Teresa L. Street, Nicholas D. Sanderson, Camille Kolenda, James Kavanagh, Hayleah Pickford, Sarah Hoosdally, Jack Cregan, Carol Taunt, Emma Jones, Sarah Oakley, Bridget L. Atkins, Maria Dudareva, Martin A. McNally, Justin O’Grady, Derrick W. Crook, David W. Eyre

**Affiliations:** Nuffield Department of Medicine, University of Oxford, John Radcliffe Hospital, Oxford, United Kingdom; NIHR Oxford Biomedical Research Centre, John Radcliffe Hospital, Oxford, United Kingdom; Department of Bacteriology, Lyon University Hospital, Lyon, France; Microbiology Laboratory, John Radcliffe Hospital, Oxford University Hospitals NHS Foundation Trust, Oxford, United Kingdom; Bone Infection Unit, Nuffield Orthopaedic Centre, Oxford University Hospitals NHS Foundation Trust, Oxford, United Kingdom; Quadram Institute Bioscience, Norwich, United Kingdom; Big Data Institute, Nuffield Department of Population Health, University of Oxford, Oxford, United Kingdom

## Abstract

**Background:** Diagnosis of orthopaedic device-related infection is challenging, and causative pathogens may be difficult to culture. Metagenomic sequencing can diagnose infections without culture, but attempts to detect antimicrobial resistance (AMR) determinants using metagenomic data have been less successful. Human DNA depletion may maximise the amount of microbial DNA sequence data available for analysis.

**Methods:** Human DNA depletion by saponin was tested in 115 sonication fluid samples generated following revision arthroplasty surgery, comprising 67 where pathogens were detected by culture and 48 culture-negative samples. Metagenomic sequencing was performed on the Oxford Nanopore Technologies GridION platform. Filtering thresholds for detection of true species versus contamination or taxonomic misclassification were determined. Mobile and chromosomal genetic AMR determinants were identified in *Staphylococcus aureus*-positive samples.

**Results:** Of 114 samples generating sequence data, species-level sensitivity of metagenomic sequencing was 49/65 (75%; 95%CI 63-85%) and specificity 103/114 (90%; 95%CI 83-95%) compared with culture. Saponin treatment reduced the proportion of human bases sequenced in comparison to 5µm filtration from a median (IQR) 98.1% (87.0%-99.9%) to 11.9% (0.4%-67.0%), improving reference genome coverage at 10-fold depth from 18.7% (0.30%-85.7%) to 84.3% (12.9%-93.8%). Metagenomic sequencing predicted 13/15 (87%) resistant and 74/74 (100%) susceptible phenotypes where sufficient data were available for analysis.

**Conclusions:** Metagenomic nanopore sequencing coupled with human DNA depletion has the potential to detect AMR in addition to species detection in orthopaedic device-related infection. Further work is required to develop pathogen-agnostic human DNA depletion methods, improving AMR determinant detection and allowing its application to other infection types.

## Introduction

Infection is a serious and challenging complication of orthopaedic-implant surgery, occurring in up to 2% of joint replacement procedures[1,2]. It may result in multiple surgeries and long-term antimicrobial treatment, with significant impacts on patient well-being and healthcare costs[1–3]. The current gold-standard for diagnosis of prosthetic joint infection (PJI) is culture of multiple peri-prosthetic tissue (PPT) samples[4–6], although this is slow and relatively insensitive, identifying a causative organism in as few as 65% of cases[4,7]. Sonication fluid culture may improve sensitivity[8] and is used alongside PPT culture in some centres[7]. More comprehensive and rapid PJI diagnostics would potentially allow earlier and more targeted treatment.

Several studies[9–15] have shown metagenomic sequencing can identify causative pathogens in PJI, achieving species-level sensitivity of 83-96%, and potentially identifying difficult to grow organisms and pathogens following prior antibiotics. However, lack of comprehensive antimicrobial susceptibility prediction from orthopaedic metagenomic sequencing has limited its application. We[16] and other authors[17–19] have applied long-read sequencing to link antimicrobial resistance (AMR) determinants to individual species within metagenomic samples. However, these approaches require extraction of sufficient pathogen DNA from clinical samples, which is often difficult given the overwhelming amount of human DNA frequently present. Without improved pathogen DNA yields only species identification is possible and AMR prediction remains challenging.

We build on previous proof-of-principle work applying Oxford Nanopore sequencing for the diagnosis of PJI from sonication fluid[12], to evaluate a laboratory protocol for human DNA removal and assess its impact on metagenomic sequencing-based AMR identification, specifically in *Staphylococcus aureus* as an example organism frequently causing PJI.

## Methods

### Sample collection and processing

Samples were collected at the Nuffield Orthopaedic Centre (NOC), a specialist musculoskeletal hospital with a dedicated Bone Infection Unit within Oxford University Hospitals (OUH). Samples collected intraoperatively from revision arthroplasty surgery undertaken for suspected infection and aseptic failure between 15-June-2018 and 22-January-2020 were obtained following routine diagnostic work-up. Use of samples and linked de-identified metadata in this study was reviewed by an NHS research ethics committee (London – Queen Square Research Ethics Committee, reference 17/LO/1420) and ethical approval was given.

Sonication fluids were generated from explanted prosthetic joints and other metalwork. Aerobic and anaerobic culture of sonication fluid and periprosthetic tissue (PPT) samples was performed as previously described[10,20]. Cultured organisms were identified using MALDI-TOF MS (Bruker, Coventry, UK). Antimicrobial susceptibility of cultured bacteria was performed according to EUCAST methods on either a BD Phoenix 100 Automated Microbiology System (Becton Dickinson, Wokingham, UK) or manually by disc diffusion. Tissue samples also underwent histological examination.

### Sample preparation, sequencing and data analysis

Details of sample preparation (including human DNA removal with saponin, DNA extraction, library preparation), nanopore sequencing, metagenomic data processing and analysis, and thresholds for taxonomic classification are provided in the supplementary material. When evaluating the specificity of species detection, species present in PPT cultures, but not sonication fluid cultures, were not considered false-positive results.

### Detecting *S. aureus* AMR determinants

AMR determinants were detected using methods previously described[16]. Briefly, chromosomal variants were called after aligning reads to a reference genome (MRSA252) using minimap2[21] (version 2.17-r941). Variants identified using Clair[22] (git commit 54c7dd4) were filtered with a trained random-forest classifier described previously[16] then compared to a database of resistance conferring SNPs[23] (Table S1). Mobile genetic elements containing AMR genes were detected by minimap2 overlaps with a catalogue of genes adapted from previous studies[23] (Table S2). Detected genes, trimmed of surrounding sequence, were realigned to the resistance gene reference sequence. The Nextflow[24] workflow used is provided at https://gitlab.com/ModernisingMedicalMicrobiology/genericbugontworkflow.

## Results

115 sonication fluid samples from 113 patients underwent culture and metagenomic sequencing (Table S3). 49/115 (43%) were culture-positive for one (n=48) or two (n=1) organisms at >50 CFU/ml, 10 (9%) had <50 CFU/ml (6/10 with a highly pathogenic organism) and 8 (7%) were culture-positive but not quantified (including one sample with three organisms isolated). 48 (42%) samples were culture-negative, of which 13 (27%) had evidence of acute inflammation on histology. Staphylococcal species were most commonly isolated, with *Staphylococcus aureus* accounting for 15/60 (25%) of all species cultured at >50 CFU/ml in sonication fluids (Table 1). Between 2-8 samples were run per sequencing flow cell using 52 flow cells overall, with a median sequence yield per flow cell of 10.2 gigabases (IQR 4.7-13.9). Efficient demultiplexing was achieved, with a median 97% (IQR 94-98%) of sequence bases assigned to a sample. Sequence data have been deposited in the European Nucleotide Archive (PRJEB42910).

**Table 1.**
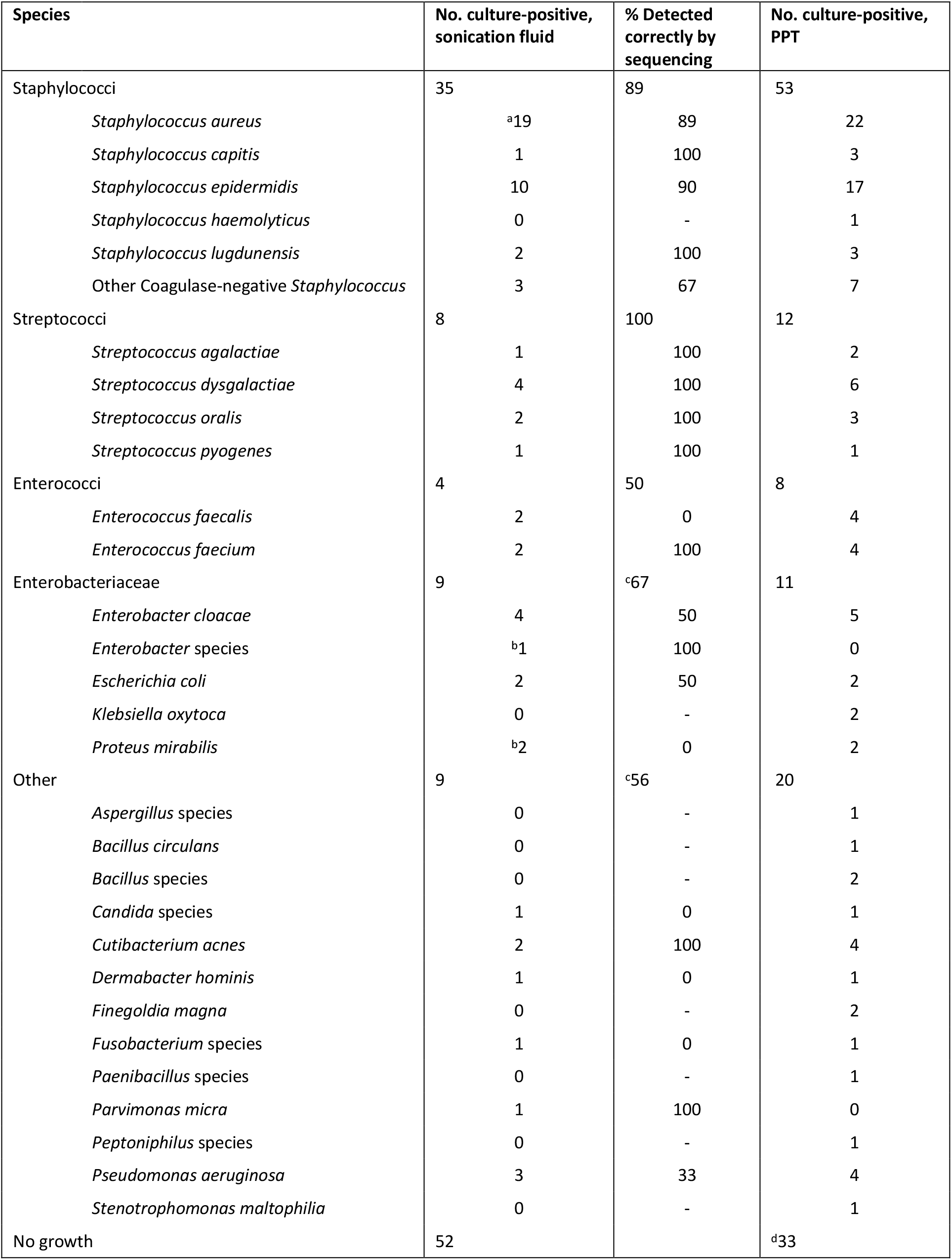
Summary of species observed in culture for sonication fluid and peri-prosthetic tissues (PPT) for 114 samples successfully sequenced. Results are reported where ≥ 1 isolate of a species was cultured. Sonication fluids were considered culture positive if > 50 CFU/ml was isolated or if <50 CFU/ml of a pathogenic organism (i.e., not skin flora) was isolated and negative if no growth or < 5 CFU/ml of an organism was isolated. ^a^includes 4 samples where S. aureus was isolated at <50 CFU/ml; ^b^includes one sample where indicated species was isolated at <50 CFU/ml; ^c^includes one species detected by sequencing to genus-level only; ^d^includes one patient where no tissue samples were collected during surgery.

### Effect of saponin on human and bacterial sequence yields

We compared human cell DNA depletion by saponin treatment to 5 µm filtration prior to DNA extraction. 91 sonication fluid samples had sufficient volume (≥80 ml) to compare both depletion methods, of which 49 were culture-positive. Treatment with 5% saponin reduced the proportion of human bases sequenced compared to 5 µm filtration (Figure 1A), from a median (IQR) 98.1% (87.0%-99.9%) to 11.9% (0.4%-67.0%) (Wilcoxon p<0.001). A >80% reduction in human bases was observed in 30/49 samples (Table S4). There was a corresponding increase in median bacterial bases sequenced from 3.1×10^7^ (IQR 5.3×10^6^-1.2×10^8^) to 3.3×10^8^ (2.2×10^7^-1.6×10^9^) (Figure 1B). Comparing the proportion of reference genome covered at 10-fold depth (indicative of sufficient data for AMR prediction), saponin treatment increased the median (IQR) from 18.7% (0.30%-85.7%) to 84.3% (12.9%-93.8%) (Figure 1C), as well as increasing genome coverage depth (Figure 1D).

**Figure 1.**
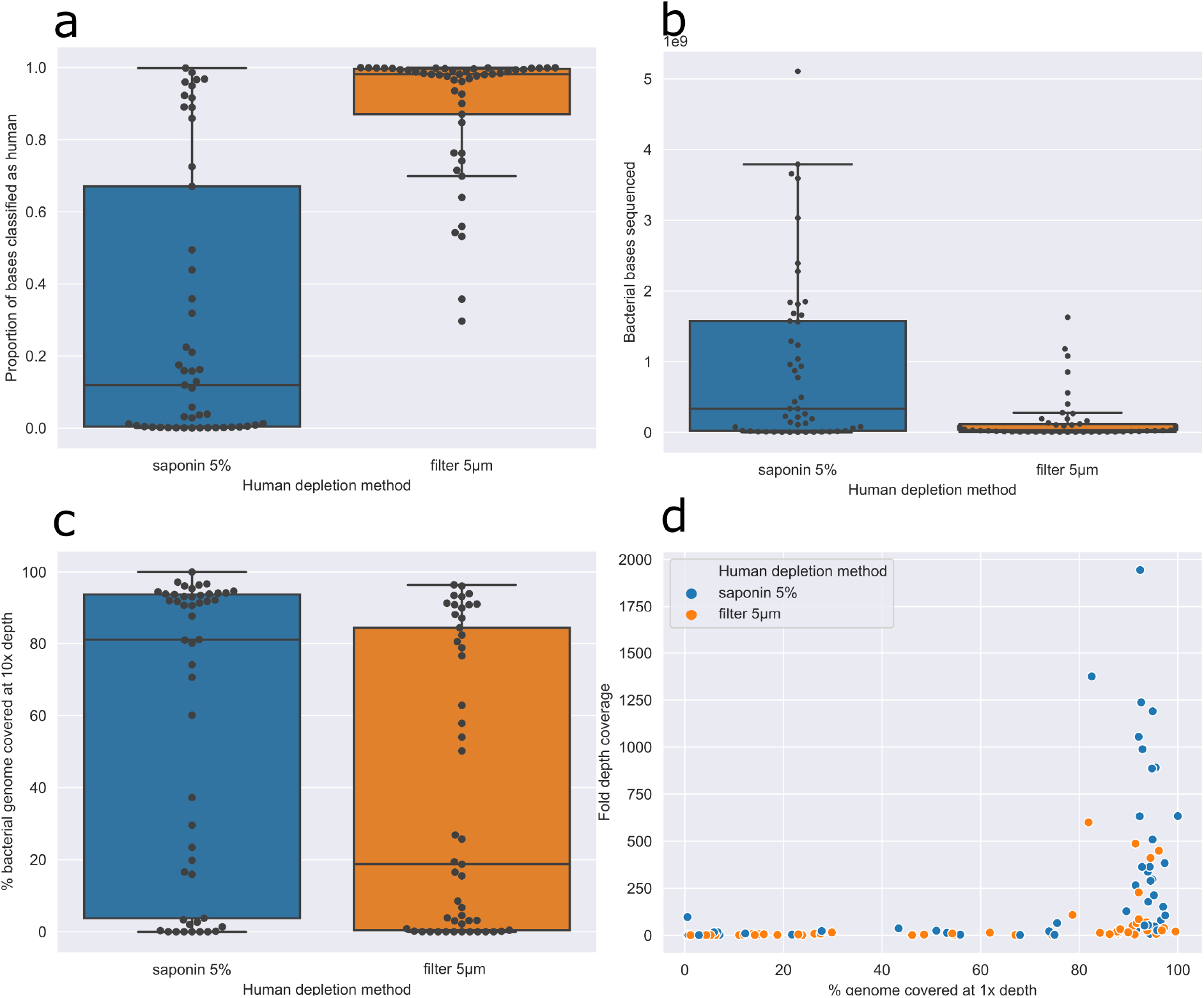
Effect of saponin treatment on human DNA depletion. Panel A, Proportion of bases sequenced classified as human by treatment used. Statistical significance determined using a paired Wilcoxon signed-rank test with a P value of 1.11×10^−9^. Panel B, Number of bacterial bases sequenced by treatment used. Effect of saponin treatment on reference genome mapping breadth (panel C) and depth (panel D).

### Species detection

Species were identified using metagenomic data from 5% saponin treated samples. One culture-positive (sample 105, positive for a single organism) failed to generate any sequence data so was excluded, leaving 114 samples in the analysis. We used filtering to remove taxonomic misclassification and contamination with thresholds determined by maximising the Youden index (sensitivity+specificity-1) (Figure 2A). We required coverage of >45% of the reference genome, or for samples with less bacterial DNA sequence: >60% of the total bacterial bases from the same species, ≥700 reads from that species with >40% of bases within the reads mapped to the reference genome (Figure 2B).

**Figure 2.**
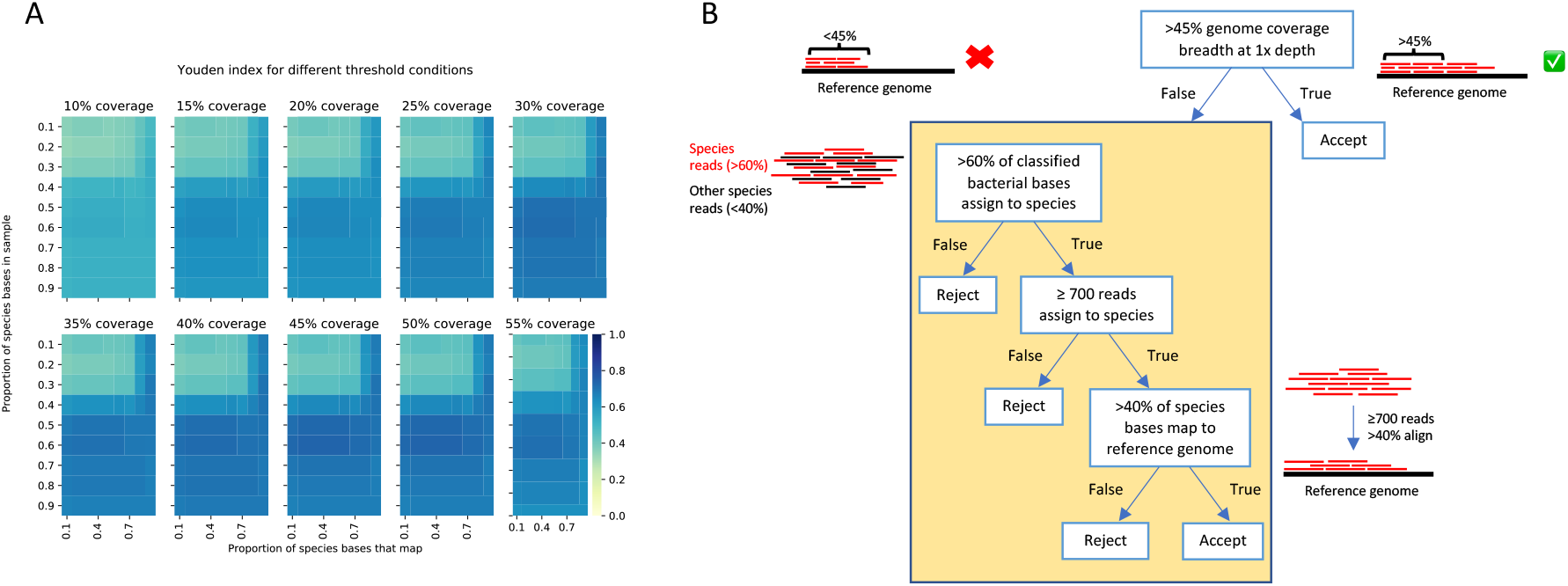
Species detection. Heatmap of Youden index (sensitivity + specificity – 1) values for each coverage breadth (panels), with proportion of species bases in the sample (y-axis) over the proportion of these bases that map to a reference genome (x-axis). Colour represents Youden index, where darker blue indicates a more optimum threshold condition (A). Final filtering selection criteria used to determine true presence of a species from metagenomic sequencing reads (B).

We used sequencing to attempt to identify 65 individual species from culture-positive sonication fluid samples (49 organisms at >50 CFU/ml, 6 highly pathogenic organisms at <50 CFU/ml and 10 unquantified by culture). Species-level sensitivity was 49/65 (75%, 95%CI 63-85%) (Table S3). Two organisms were identified to genus-level only by sequencing; hence the genus-level sensitivity was 51/65 (78%, 67-88%). Of the 14 species cultured and not identified by sequencing, 11 were present but below filtering thresholds (4 cultured at >500 CFU/ml, 4 cultured at 50-500 CFU/ml, 2 were pathogenic organisms at < 50 CFU/ml and 1 was not quantified in culture) and 3 (two *Pseudomonas aeruginosa* and one *Candida parapsilosis*) were not detected in sequence data in samples treated with 5% saponin.

Specificity, defined as no ‘false-positive’ species per sample compared to sonication fluid culture, was 103/114 (90%, 95%CI 83-95%) (Table S3). Considering only the 48 culture-negative samples, specificity was 47/48 (98%, 89-100%). In 11 samples (10 with other species identified) 16 additional species were detected by sequencing. These species included five likely bioinformatic misclassifications leading to identification of an additional species from the same genus: 3 additional *En*t*erobacter cloacae* complex species where sonication fluid cultured *E. cloacae* and one extra *Dermabacter* and *Streptococcus* species. Five species were plausible anaerobic infections (*Fusobacterium nucleatum, Prevotella intermedia, Cutibacterium acnes* and *Anaerococcus mediterraneensis*); and two had skin commensal species also cultured in sonication fluid at <50 CFU/ml but not in corresponding PPT (*Staphylococcus epidermidis*). Four samples had other additional species not otherwise identified by culture (Table S5), including one sample that initially appeared to show potential sample-to-sample cross-contamination, detecting *S. aureus* in a culture-negative sonication fluid (sample 53) sequenced in the same batch as a *S. aureus*-positive sample (sample 54). However, the two sequences were >10,000 SNPs different (Figure S1), suggestive, along with evidence of acute inflammation on histology, of true infection. In two patients, two separate sonication fluids were cultured from the same surgery. Sequencing matched culture results in both samples for one patient (culture-negative and no species detected by sequencing, samples 128a and 128b). For the other patient, although one sonication fluid was culture-positive for *Streptococcus dysgalactiae* and the other negative, 4/5 corresponding PPT samples grew *S. dysgalactiae* and both sonication fluid sequences identified *S. dysgalactiae* (samples 40a and 40b).

Only 1/45 negative controls contained a species above the filtering thresholds: *S. aureus*. The control was treated with saponin and extracted in the same batch as a *S. aureus* culture-positive sample. It was not possible to include this sequence in the *S. aureus* phylogenetic tree (Figure S1) due to insufficient data.

### AMR prediction for *Staphylococcus aureus*

Sequencing detected *S. aureus* in 17/19 *S. aureus* culture-positive sonication fluids and in 2 sonication fluid culture-negative samples with *S. aureus* culture-positive PPT. 14/19 had a single-fold genome coverage >90% (Table 2). For all 19 samples, we compared sequencing-based AMR predictions with phenotypic results for 8 antimicrobials.

**Table 2.**
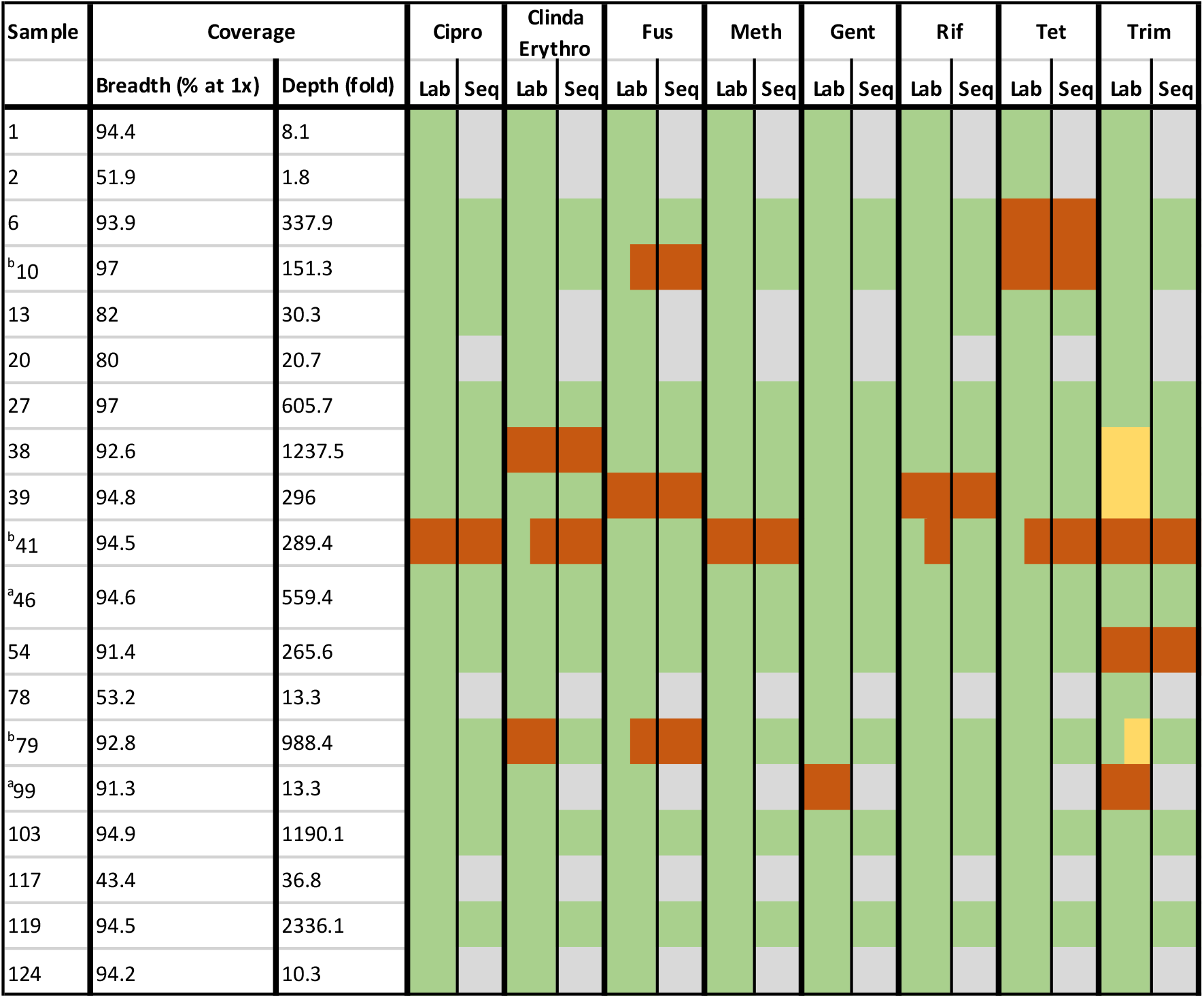
*Staphylococcus aureus* genome coverage, lab sensitivities, and predicted sensitivities from sequencing. Green, Sensitive; Orange, Resistant; Yellow, Intermediate; Grey, Unable to determine due to lack of coverage. Cipro, ciprofloxacin; Cinda, clindamycin; Erythro, erythromycin; Fus, fusidic acid; Meth, methicillin; gent, gentamicin; Rif, rifampicin; Tet, tetracycline; Trim, trimethoprim. ^a^Samples 46 and 99 were culture-negative sonication fluids, results reported here are from corresponding culture-positive PPT for comparison purposes. ^b^Sensitivities reported for both *S. aureus* morphotypes cultured in these samples.

Across 152 organism-drug combinations, sequencing correctly predicted susceptibilities in 87/152 (57%), was unable to call susceptibilities in 60/152 (39%) due to insufficient data, and made an incorrect prediction in 5/152 (3%) (two very major errors [missed resistance], three minor errors [intermediate phenotype called susceptible], Table S6). In three cases 2 morphotypes were reported with differing sensitivities in the laboratory (samples 10, 41, and 79: with differing sensitivities for fusidic acid [samples 10 and 79], trimethoprim [sample 79] and for clindamycin/erythromycin, rifampicin and tetracycline [sample 41]). Counting both susceptibilities as a single combination, sequencing was reported as correct if the resistant genotype was detected.

Sequencing correctly predicted 13/17 (76%) resistance phenotypes reported by the laboratory, with insufficient data for prediction in 2 (12%) and an incorrect result in 2 (12%, described below). Sequencing correctly predicted 74/74 (100%) susceptible phenotypes where sufficient data were available, with insufficient data for 58 susceptible organism-drug combinations. In 3 cases where the laboratory reported an intermediate sensitivity to trimethoprim (samples 38, 39 and 1 of 2 morphotypes in sample 79) sequencing predicted susceptible.

In the 8 cases where a successful prediction of antimicrobial susceptibility was across all drugs, a combination of high genome coverage breadth and depth was observed (Table S6) (>92% breadth at >151-fold depth). Genome breadth or depth of coverage was insufficient in the 5 cases where a partial prediction was made: where breadth was >90%, depth was ≤10-fold, or where depth was >10-fold, breadth was <80%.

We correctly identified the only methicillin-resistant *S. aureus* (sample 41; 54-fold coverage of *mecA*). Sequencing also correctly identified the only ciprofloxacin-resistant *S. aureus* (sample 41, *grlA* S80F and *gyrA* S84L). We detected rifampicin resistance in sample 39 (*rpoB* A477V), and for sample 41, which yielded rifampicin sensitive and resistant isolates, sequencing detected a C→T mutation in 20% of reads mapping to *rpoB* conferring the A477V substitution (Figure S2). Our approach, based on the sequence of the majority of reads, called this sample as susceptible, missing the resistant isolate as our approach does not currently account for mixed populations within the same species.

The second major error identified involved failure to detect any resistance determinants for clindamycin/erythromycin in sample 79, despite high genome coverage (93% at an average depth of 988-fold) despite a resistant phenotype for these agents.

Only 3 S. *aureus* culture-positive samples not treated with saponin prior to DNA extraction (samples 6, 78 and 79) achieved enough breadth and depth of genome coverage to successfully detect any AMR determinants, with predictions made for ciprofloxacin, rifampicin and fusidic acid only.

## Discussion

Here we describe the largest study to date of long-read metagenomic sequencing applied to PJI. We show that saponin can effectively deplete human DNA. Using samples treated with saponin the sensitivity and specificity for species-level detection was 75% and 90%, respectively. Using *Staphylococcus aureus* as an exemplar organism, we successfully predicted 13/15 (87%) resistant and 74/74 (100%) susceptible phenotypes where sufficient sequenced data was available; 60/152 (39%) drug-organism combinations lacked sufficient data. 5/152 (3%) yielded incorrect results (three minor errors, intermediate isolates called susceptible and two very major errors, resistant isolates called sensitive).

Saponin has previously been shown to reduce host cell contamination in respiratory samples and CSF[17,25,26]; here in orthopaedic infection-related samples saponin decreased the proportion of human DNA sequenced from a median of 98% to 12%, with an increase in bacterial genome coverage and allowing the detection of AMR determinants in *S. aureus* culture-positive samples. However, 11/49 samples compared achieved incomplete depletion with a <12% reduction in human bases. Although confirmation of successful human DNA depletion could be performed by qPCR, identifying cases where a second depletion might be useful prior to sequencing, this would make workflows more burdensome.

While saponin treatment enhanced recovery of bacterial DNA overall, it appeared to adversely affect some species. *Enterococcus faecalis* was not identified by sequencing in two culture-positive samples. While it was detected well below the filtering thresholds in both cases, for one of these two samples, with a paired 5μm filter sample, the number of reads classified as *E. faecalis* was much higher without saponin treatment. Similar results were observed in 2 of 3 samples culture-positive for *Pseudomonas aeruginosa*, where sequencing reads were only detected in the 5μm filter samples and not after saponin treatment. We hypothesise that sample storage at 4°C between collection and treatment could have an adverse effect on the extraction of bacterial DNA from some species.

Taxonomic misclassification explained the failure to detect *Dermabacter hominis* found on culture, where sequencing identified the wrong species, *D. vaginalis*, as *D. hominis* was not present in the species classification database. We also failed to identify a *Candida parapsilosis* infection, as the lysis step of our DNA extraction protocol was not designed to disrupt fungi. Sequencing also failed to identify to species level above the filtering thresholds ten remaining pathogens seen in culture.

Sequencing identified additional species not detected by culture of sonication fluid in 11 cases. In some instances, this may represent taxonomic misclassification of similar species, e.g. within the *Enterobacter cloacae* complex or between *Streptococcus* species. We also identified three anaerobic organisms not detected by standard anaerobic culture: *Fusobacterium nucleatum*, a known cause of PJI[27], *Prevotella intermedia*, previously reported in a case of osteomyelitis[28], and *Anaerococcus mediterraneensis. Cutibacterium acnes* was identified in two samples. *C. acnes* is a common contaminant, but also a well-known cause of PJI. We observed that in *C. acnes* culture-positive samples the number of bases sequenced, along with genome breadth and depth of coverage, are very high: 100% and 96% breadth with 633-fold and 448-fold depth of coverage, respectively, in the two *C. acnes*-positive samples (samples 12 and 58). One “false-positive” *C. acnes* had a genome coverage breadth and depth of 97% and 384-fold, similar in magnitude to the true positives, i.e. more suggestive of true infection than contamination. These results support our previous observation that species-specific filtering may be useful in the case of *C. acnes*[10] but larger sample numbers are required to address this.

The importance of including negative controls for metagenomic sequencing is widely recognised. Laboratory reagents are known to contribute to low level contamination[29,30] and previous studies recognise the perils of misinterpreting contamination for true clinical findings[31,32]. Sequencing identified one species in one of our 45 negative controls. However, these findings depended on filtering our data; while we were able to filter out low-level contaminating DNA and identify most true infections, this likely reduced sensitivity and further independent studies are needed to validate the filtering thresholds chosen.

In the case of *Staphylococcus aureus* infection, we were able to successfully predict antimicrobial resistance in 87% and susceptibility in 100% of cases where sufficient sequence data was available. Of the 152 possible organism-drug combinations, insufficient data for prediction was obtained in 39%, highlighting the need for further optimisation around laboratory protocols. In 6 cases where two different morphotypes of *S. aureus* displayed differing sensitivities, sequencing was able to successfully predict resistance in 4 cases. Resistance determinants were detected in a proportion of sequence reads in the fifth case, serving to emphasise the need for bioinformatic methods that account for mixed infections. The sixth case with both sensitive and intermediate susceptibility to trimethoprim, was reported by sequencing as sensitive (as were 2 other samples that were reported with an intermediate phenotype).

Our work adds to other recent reports. Wang *et al*[13] assessed nanopore sequencing for diagnosis of PJI, identifying the cause of infection in 4/5 sonication fluid samples. A small number of AMR determinants were identified in 2/5 samples, although concordance with laboratory sensitivities was variable. Yan *et al*[33] previously assessed a commercial metagenomic data analysis service for the detection of antimicrobial resistance determinants specifically in Staphylococcal species from sonication fluid and reported sensitivities ranging from 66-85%, similar to the 87% we observed in this study. Metagenomic sequencing for AMR determinant detection has also been applied to PPT samples for bone and joint infections[9], with the correct susceptibility inferred in 94.1% of monomicrobial and 76.5% of polymicrobial samples. Recent reviews also highlight the use of metagenomic sequencing for AMR detection[34,35].

Limitations of metagenomic sequencing, observed both here and in previous studies (e.g.[11,36]) include the need for each species to be present in the reference database, and also the need for unbiased pan-bacterial/pan-fungal species-agnostic DNA extraction (e.g. here we fail to detect a *Candida* species on this basis). The need to filter data means that low-level and polymicrobial infections may be erroneously filtered-out and missed. Finally, successful AMR prediction relies on obtaining a combination of high genome coverage breadth and depth, which is directly linked to the proportion of human DNA sequenced.

In conclusion, metagenomic sequencing is a useful addition to the repertoire of diagnostic tests performed in the case of suspected PJI. Although sensitivity remains below that which would be needed to replace culture as a gold standard diagnostic method, the detection of additional pathogens is useful in culture-negative samples. Saponin is a useful method for host DNA depletion, but adverse effects on some pathogens mean that better approaches to human DNA depletion that preserve all microbial DNA are required. Additionally, a microbial DNA extraction method that encompassed all pathogens is necessary so fungal infections can be detected in addition to bacterial. Our study has demonstrated as a proof of principle that nanopore sequencing in conjunction with human DNA depletion shows potential for the detection of AMR determinants using *S. aureus* as a model organism for PJI. Further development is required to test this method on a wider range of pathogens.

## Data Availability

Sequence data have been deposited in the European Nucleotide Archive (PRJEB42910)

## Data Availability

Sequence data have been deposited in the European Nucleotide Archive (PRJEB42910)

## Acknowledgements

We thank the microbiology laboratory staff of the John Radcliffe Hospital, Oxford University Hospitals NHS Trust, for providing assistance with sample collection and processing.

