## Supplementary Material for "Clinical Metagenomic Sequencing for Species Identification and Antimicrobial Resistance Prediction in Orthopaedic Device Infection"

#### Supplementary Methods

##### Human DNA depletion and bacterial DNA extraction

We tested the impact of host cell DNA depletion with saponin, using the method outlined by Charalampous et al[1]. Prior to human DNA depletion and bacterial DNA extraction, 40 ml of fresh sonication fluid, stored at 4°C between generation and use, was concentrated by centrifugation as previously described[2], yielding an initial volume of 1-2 ml. After further centrifugation at 8000 g for 5 minutes, the pellet was resuspended in 250 µl of PBS and 200 µl of 5% saponin solution was added, giving a final concentration of 2.2% in each depletion reaction. As a control, an equal volume of sonication fluid was concentrated by centrifugation and passed through a 5 µm syringe filter instead of treatment with saponin. After host DNA depletion, bacterial DNA was extracted by mechanical lysis followed by ethanol precipitation, as previously described[2]. DNA was purified using AMPure XP solid phase reversible immobilisation (SPRI) beads (Beckman Coulter, High Wycombe, UK) and eluted in a final volume of 26 µl TE buffer. Negative controls for sonication (0.9% saline) and before saponin treatment (PBS) or 5 µm filtration (0.9% saline) were prepared alongside the extractions from sonication fluids.

##### Library preparation and sequencing

DNA extracts were prepared for sequencing on an Oxford Nanopore Technologies (ONT, Oxford, UK) GridION using the Rapid PCR Barcoding Kit (SQK-RPB004, ONT) and a modified protocol as previously described[3]. Initially, PCR libraries were purified individually with AMPure XP beads and eluted in a total volume of 10 µl before quantification on a Qubit 2.0 fluorimeter with the Quant-iT dsDNA HS Assay kit (Life Technologies, Paisley, UK). Purified libraries were multiplexed up to a maximum of 8, with a total of between and 25 and 302 fmol of pooled library loaded per flow cell, for the first 11 flow cells.  Subsequent libraries were quantified immediately post-PCR, pooled together by similar concentrations then purified with AMPure XP beads and eluted in 10 µl. Here, a total of between 6 and 84 fmol of pooled library, corresponding to a maximum of 8 multiplexed libraries per flow cell, were loaded for all subsequent flow cells.

#### Nanopore sequence processing and analysis

Nanopore sequences were basecalled and demultiplexed using Guppy (Oxford Nanopore Technologies, Version 3.1 or higher) automatically on the GridION platform. Sequences were analysed using our in-house workflow CRuMPIT, described previously[4]. Briefly, sequences are classified with Centrifuge[5], binned into species-specific groups and aligned to a reference genome for that species. Reads classified as human are discarded and reads classified to a lower resolution taxon than species, e.g. genus only, are not mapped. Some runs were performed prior to Guppy providing demultiplexing by default on the GridIONs and in these cases Porechop v0.2.4[6] was used for demultiplexing. The fastq files from these runs had adapters trimmed and therefore could not be demultiplexed again with Guppy, so here we basecalled from fast5 files and then demultiplexed these runs with Guppy to maintain consistency with later runs. Human reads had, however, already been discarded from these fast5 files to comply with ethical requirements, so human read number comparisons use a mix of results from either Porechop or Guppy. Since comparisons are between treatments of the same sample sequenced on the same run the analysis should not be affected.

#### Determining species detection performance

Species classifications from Centrifuge and CRuMPIT mapping metrics, including coverage breadth and proportion of bases mapping to the reference, were compared to standard microbiological culture results. We used species presence in sonication fluid culture at >50 CFU/ml (or ≤50 CFU/ml of a highly pathogenic organism) as the reference standard for presence of a bacterial species. When evaluating specificity, species present in PPT cultures, but not sonication fluid cultures, were not considered false-positive results.

Where standard culture was reported to the genus level only, species belonging to the same genus were counted as a match. In this setting, multiple species matching to one genus report were reduced to a single match to avoid artificially inflated sensitivity results.

#### Species detection filtering

To distinguish between true species classified by Centrifuge and misclassifications or low-level contamination, filtering thresholds were determined. We used three metrics for filtering. The first used the percentage of the identified species reference genome covered by sequence reads. Secondly, if the sample had low overall numbers of bacterial reads with no species identified above the given percent coverage, the proportion of bases classified as a species compared to total bacterial bases in the sample was used. Thirdly, as reads can be classified with lower specificity by Centrifuge to improve sensitivity, the proportion of species bases mapping to the reference genome was also considered. Thresholds for each filter were determined by choosing the combination of thresholds that maximised the Youden index (specificity+sensitivity-1).

### Supplementary Data

#### Supplementary Figures

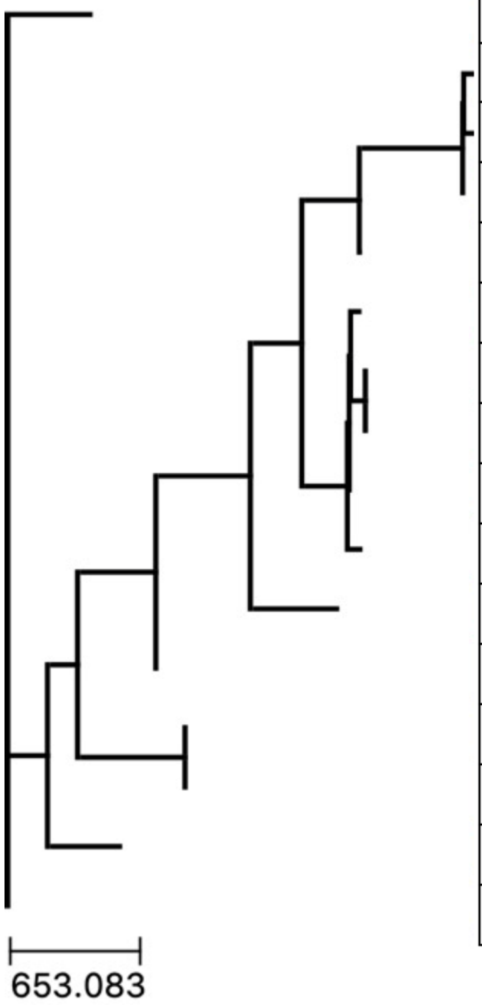


| Run number | Sample number | Human depletion method | *S. aureus* culture-positive? | # SNP differences to nearest neighbour[a] |
|---|---|---|---|---|
| 4 | 10 | Saponin 5% | TRUE | 5687 |
| 56 | 103 | Saponin 5% | TRUE | 72 |
| 63 | 119 | Saponin 5% | TRUE | 83 |
| 13 | 27 | Saponin 5% | TRUE | 72 |
| 33 | 53 | Saponin 5% | FALSE | 5178 |
| 46 | 78 | Filter 5μm | TRUE | 177 |
| 52 | 79 | Saponin 5% | TRUE | 0 |
| 52 | 79 | Filter 5μm | TRUE | 0 |
| 33 | 54 | Saponin 5% | TRUE | 298 |
| 55 | 99 | Saponin 5% | TRUE | 376 |
| 26 | 38 | Saponin 5% | TRUE | 9874 |
| 28 | 41 | Saponin 5% | TRUE | 8675 |
| 2 | 6 | Filter 5μm | TRUE | 0 |
| 2 | 6 | Saponin 5% | TRUE | 0 |
| 29 | 46 | Saponin 5% | TRUE | 5921 |
| 27 | 39 | Saponin 5% | TRUE | 5892 |

***Figure S1. Phylogenetic tree of Staphylococcus aureus genome sequences.*** *There are >10k SNPs between sequences for sample 53 and 54, and over 5k SNPs between sample 53 and other samples sequenced in the lab. [a]Dark blue represents SNP differences >5000, light blue represents SNP differences <5000.*

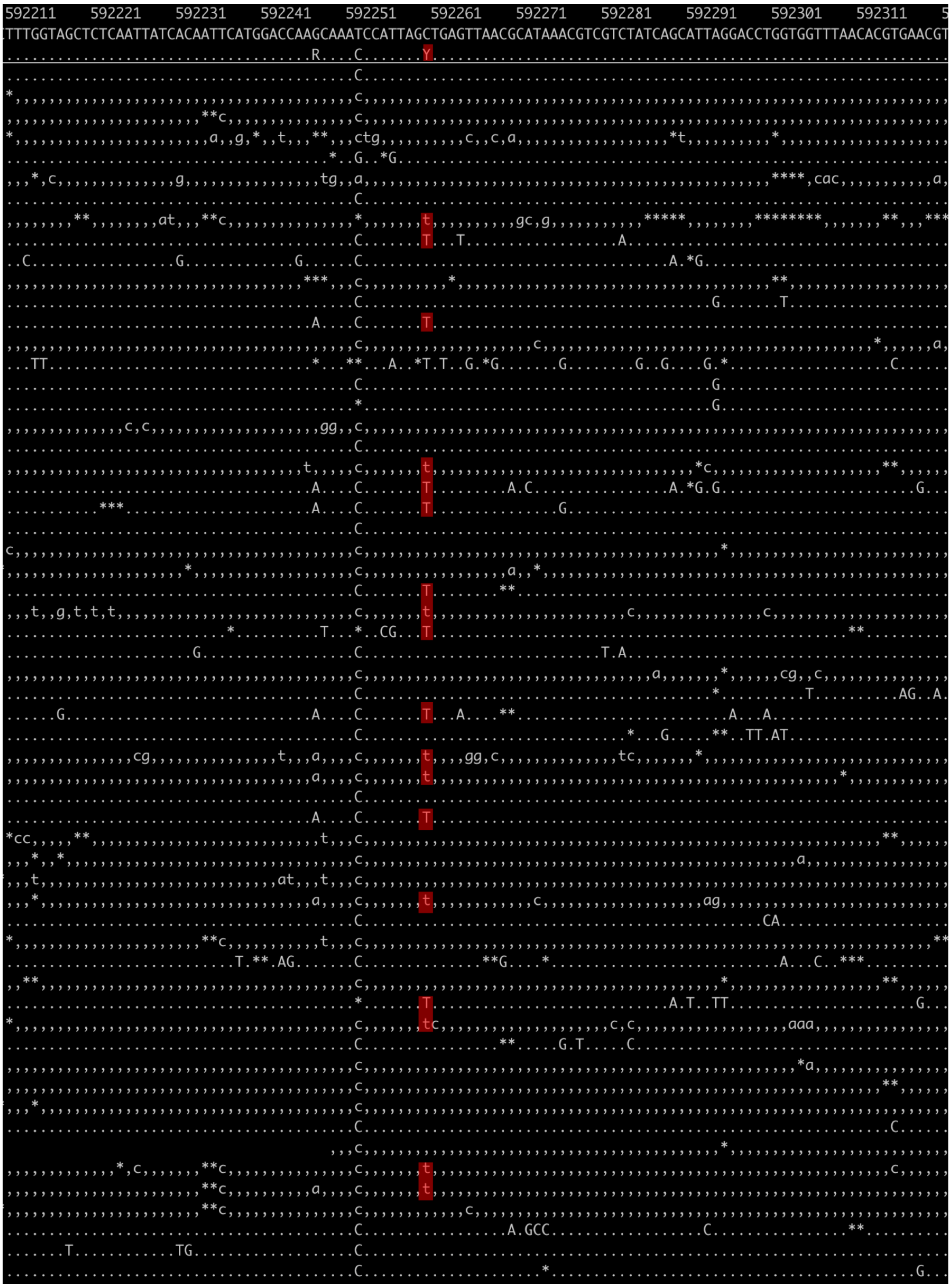

***Figure S2. rpoB mutation observed in sample 41 at position 592260 in the reference genome.***

*Nucleotide mutation C to T causing amino acid codon substitution from GCT (A) to GTT (V) at residue 477, conferring resistance to rifampicin, highlighted in red. T substitution observed in 20% of reads aligning to this position.*

### Supplementary Tables

| **Antimicrobial agent** | **Gene** | **Amino acid substitutions** | **Reference gene accession no. (nucleotide positions)** |
|---|---|---|---|
| Ciprofloxacin | *gyrA* | S84L, E88K, G106D, S85P, E88G, E88L | BX571857.1 (7005–9668) |
| | *grlA* | S80F, S80Y, E84K, E84G, E84V, D432G, Y83N, A116E, A48T, D79V, V41G, S108N | BX571857.1 (1386869–1389271) |
| | *grlB* | R470D*, E422D*, P451S*, P585S*, D443E*, R444S* | BX571857.1 (1384872–1386869) |
| Fusidic acid | *fusA* | A160V*, A376V, A655E, A655P*, A655V*, A67T*, A70V*, A71V*, B434N, C473S*, D189G*, D189V*, D373N*, D463G*, E233Q*, E444K, E444V*, E449K*, F441Y, F652S*, G451V, G452C, G452S, G556S, G617D, G664S, H438N, H457Q, H457Y, L430S*, L456F, L461K, L461S, M161I*, M453I, M651I, P114H, P404L, P404Q, P406L, P478S, Q115L, R464C, R464H, R464S, R659C, R659H, R659L, R659S, R76C*, S416F*, T385N, T387I*, T436I, T656K, V607I, V90A, V90I, Y654N* | BX571857.1 (577685–579766) |
| Rifampicin | *rpoB* | A473T*, A477D, A477T*, A477V, D471G*, D471Y, D550G, H481D, H481N, H481Y, I527F, I527L*, I527 M*, ins 475H, ins G475*, L466S*, M470T*, N474K*, Q456K, Q468K, Q468L, Q468R, Q565R*, R484H, S463P, S464P, S486L, S529L* | BX571857 (568813–572364) |
| Trimethoprim | *dfrB* | F99Y, F99S, F99I, H31N, L41F, H150R, L21V*, N60I* | BX571857.1 (1464014–1464493) |

***Table S1. Polymorphisms conferring resistance in chromosomal genes.*** *Asterisk (*) represent mutations in combination. Adapted from Gordon* et al.*[7]*

| Antimicrobial agent(s) | Gene | Product | Reference gene accession no. (nucleotide positions) |
|---|---|---|---|
| Methicillin | *mecA* | Low-affinity PBP2 | BX571856.1 (44919–46925) |
| Erythromycin | *msrA* | Erythromycin resistance protein | CP003194 (54168–55634) |
| Erythromycin and clindamycin | *ermA* | rRNA adenine N-6-methyltransferase | BA000018.3 (56002–56733) |
| | *ermB* | rRNA adenine N-6-methyltransferase | AB699882.1 (4971–5708) |
| | *ermC* | rRNA adenine N-6-methyltransferase | HE579068 (7858–8592) |
| | *ermT* | 23S rRNA methylase | HF583292 (11344–12078) |
| | *lsaE* | ATP-binding cassette (ABC) ribosomal protection protein, family F | JX560992 (11387-12872) |
| | *vgaE* | ATP-binding cassette (ABC) ribosomal protection protein, family F | FR772051 (8740-10315) |
| | *lnuA* | Lincosamide nucleotidyltransferase | AM399080 (1664-2150) |
| | *lnuB* | Lincosamide nucleotidyltransferase | AY183453.1 (2730-3950) |
| | *ereA* | Erythromycin esterase | X03988.1 (383-1642 |
| | *ereB* | Erythromycin esterase | AE007317.1 (383-1642) |
| | *mefE* | Macrolide efflux pump | FJ196385.1 (11084-12313) |
| | *mefB* | Macrolide efflux pump | AB571865.1 (144313-145536) |
| | *mefC* | Macrolide efflux pump | MN728681.1 (17459-18658) |
| | *mefD* | Macrolide efflux pump | AB013298.1 (2296-3195) |
| | *mphC* | Macrolide phosphotransferase | AJ238249.1 (127-930) |
| Tetracycline | *tetK* | MFS tetracycline efflux pump | FN433596 (69118–70497) |
| | *tetL* | MFS tetracycline efflux pump | HF583292 (7713–9089) |
| | *tetM* | Ribosomal protection protein | CP002643 (427033–428952) |
| Fusidic acid | *fusB* | Fusidic acid detoxification | CP003193.1 (1336–1977) |
| | *fusC* | Fusidic acid detoxification | BX571857.1 (52820-53458) |
| | *far* | Ribosome protection protein | AY373761.1 (19072–19713) |
| Trimethoprim | *dfrA* | Insensitive dihydrofolate reductase | CP002120 (2093303–2093788) |
| Trimethoprim | *dfrG* | Insensitive dihydrofolate reductase | FN433596 (502263–502760) |
| Gentamicin | *aacA-aphD* | 6′-aminoglycoside N-acetyltransferase/2″-aminoglycoside phosphotransferase | FN433596.1 (2209531–2210970) |

***Table S2. Mobile resistance genes.*** *Presence of these genes is associated with resistance to the respective antibiotic. Adapted from Gordon* et al.*[7]*

| Sample | Saponin human proportion | Filter human proportion | % of original human proportion | % reduction in human proportion of bases |
|---|---|---|---|---|
| 1 | 0.97 | 0.98 | 98.66 | 1.34 |
| 6 | 0.00 | 0.72 | 0.28 | 99.72 |
| 8 | 0.99 | 0.99 | 99.82 | 0.18 |
| 9 | 0.03 | 1.00 | 2.85 | 97.15 |
| 10 | 0.01 | 0.99 | 1.29 | 98.71 |
| 12 | 0.13 | 0.98 | 13.19 | 86.81 |
| 18 | 0.01 | 0.93 | 0.60 | 99.40 |
| 20 | 0.00 | 0.74 | 0.16 | 99.84 |
| 22 | 0.00 | 0.70 | 0.51 | 99.49 |
| 26 | 0.32 | 0.98 | 32.51 | 67.49 |
| 31 | 0.89 | 1.00 | 88.98 | 11.02 |
| 33 | 0.00 | 0.96 | 0.29 | 99.71 |
| 34 | 0.67 | 1.00 | 67.10 | 32.90 |
| 38 | 0.00 | 0.99 | 0.46 | 99.54 |
| 39 | 0.04 | 1.00 | 3.67 | 96.33 |
| 41 | 0.00 | 0.76 | 0.12 | 99.88 |
| 42 | 0.16 | 0.85 | 18.68 | 81.32 |
| 43 | 0.01 | 0.36 | 2.00 | 98.00 |
| 45 | 0.18 | 0.99 | 17.65 | 82.35 |
| 54 | 0.03 | 0.98 | 3.23 | 96.77 |
| 55 | 0.12 | 1.00 | 11.95 | 88.05 |
| 56 | 0.89 | 0.99 | 90.20 | 9.80 |
| 57 | 0.49 | 1.00 | 49.49 | 50.51 |
| 58 | 0.04 | 0.76 | 5.10 | 94.90 |
| 59 | 0.21 | 0.98 | 21.41 | 78.59 |
| 60 | 0.00 | 0.64 | 0.46 | 99.54 |
| 61 | 0.00 | 0.56 | 0.12 | 99.88 |
| 62 | 0.16 | 1.00 | 16.27 | 83.73 |
| 63 | 0.00 | 0.54 | 0.24 | 99.76 |
| 66 | 0.36 | 1.00 | 35.99 | 64.01 |
| 72 | 0.00 | 0.97 | 0.03 | 99.97 |
| 74 | 0.44 | 1.00 | 43.95 | 56.05 |
| 75 | 0.72 | 0.99 | 73.10 | 26.90 |
| 78 | 0.11 | 0.97 | 11.51 | 88.49 |
| 79 | 0.00 | 0.87 | 0.10 | 99.90 |
| 81 | 0.00 | 0.53 | 0.12 | 99.88 |
| 87 | 0.92 | 1.00 | 92.61 | 7.39 |
| 90 | 1.00 | 1.00 | 99.93 | 0.07 |
| 96 | 0.96 | 1.00 | 96.15 | 3.85 |
| 100 | 0.86 | 0.94 | 91.84 | 8.16 |
| 103 | 0.01 | 0.99 | 1.16 | 98.84 |

| | | | | |
|---|---|---|---|---|
| 104 | 0.06 | 0.98 | 5.94 | 94.06 |
| 109 | 0.95 | 1.00 | 95.17 | 4.83 |
| 112 | 0.00 | 0.90 | 0.19 | 99.81 |
| 115 | 0.22 | 0.30 | 75.68 | 24.32 |
| 116 | 0.01 | 0.98 | 0.89 | 99.11 |
| 117 | 0.16 | 0.99 | 16.04 | 83.96 |
| 120 | 0.92 | 0.98 | 93.38 | 6.62 |
| 126 | 0.97 | 0.98 | 98.35 | 1.65 |

*Table S4. Effect of 5% saponin treatment on proportion of human bases sequenced. Percent reduction in proportion of human bases sequenced following saponin treatment in comparison to 5µm filter treatment. 100% reduction means all human bases removed.*

| Sample | False-positive species | Evidence of acute inflammation on histology? | Ref. genome coverage (%) | Average depth (fold) | Interpretation |
|---|---|---|---|---|---|
| 3 | *Enterobacter ludwigii* | Yes | 54 | 3 | [a]*Enterobacter cloacae* complex species |
| | *Cutibacterium acnes* | | 59 | 2 | Plausible anaerobic pathogen/skin flora contamination |
| 20 | *Fusobacterium nucleatum* | Yes | 79 | 363 | Plausible anaerobic pathogen |
| | *Escherichia coli* | | 48 | 6 | Laboratory contamination |
| 25 | *Enterobacter hormaechei* | Yes | 92 | 628 | [a]*Enterobacter cloacae* complex species |
| 53 | *Staphylococcus aureus* | Yes | 89 | 6 | Sample-to-sample contamination/plausible infection |
| 59 | *Corynebacterium segmentosum* | Yes | 74 | 38 | Skin flora contamination/plausible infection |
| 81 | *Streptococcus* sp. NPS 308 | Yes | 67 | 121 | Correct to genus level/[b]misclassification |
| 99 | *Dermabacter vaginalis* | Yes | 58 | 2.5 | Correct to genus level |
| | *Anaerococcus mediterraneensis* | | 62 | 100 | [b]Misclassification/plausible anaerobe |
| | *Prevotella intermedia* | | 90 | 40 | Plausible anaerobe |
| | *Enterobacter hormaechei* | | 87 | 26 | [a]*Enterobacter cloacae* complex species |
| 109 | *Staphylococcus epidermidis* | Yes | 93 | 52 | Cultured at <50 CFU/ml in sonication fluid |
| 112 | *Cutibacterium acnes* | Yes | 97 | 383 | Plausible anaerobic pathogen/skin flora contamination |
| 116 | *Staphylococcus epidermidis* | Yes | 78 | 15 | Cultured at <50 CFU/ml in sonication fluid |
| 120 | *Corynebacterium segmentosum* | No | 72 | 11 | Skin flora contamination/plausible infection |

***Table S5. Additional species identified.*** *Summary of potential 'false-positive' species identified by metagenomic sequencing, showing percentage of reference genome mapped and the average fold-depth of coverage. [a]Enterobacter cloacae complex species, indicates where E. cloacae was observed by culture in these samples; [b]misclassification, suggestive of bioinformatic misclassification to a different species of the same genus.*

| Sample | % coverage of mobile gene | | | | Fold-coverage of chromosomal genes | | | | | | | | Detected gene mutations in chromosomal genes | | | | | |
|---|---|---|---|---|---|---|---|---|---|---|---|---|---|---|---|---|---|---|
| | *ermA* | *ermC* | *fusC* | *tetK* | *blaZ* | *dfrB* | *fusA* | *grlA* | *grlB* | *gyrA* | *mecA* | *rpoB* | *dfrB* | *fusA* | *grlA* | *grlB* | *gyrA* | *rpoB* |
| 1 | N | N | N | N | 56.2 | 3.6 | 12.9 | 5.3 | 7.9 | 13.2 | N | 9.2 | N | B434D, E449I, F441T, M161P, T387D | N | R470F, E422N, P451I, P585M, D443G | N | Q456S, Q565A |
| 6 | N | N | N | 100 | 1122.4 | 90.2 | 83.5 | 144.9 | 166.8 | 124.3 | N | 89.0 | N | B434D, E449I, F441T, M161P, T387D | N | R470F, E422N, P451I, P585M, D443G | N | Q456S, Q565A |
| 10 | N | N | 100 | 100 | 326.6 | 45.6 | 100.2 | 80.9 | 96.3 | 120.9 | N | 129.0 | N | B434D, E449I, F441T, M161P, T387D | N | R470F, E422N, P451I, P585M, D443G | N | Q456S, Q565A |
| 13 | N | N | N | N | 141.6 | 2.0 | 34.2 | 83.0 | 51.1 | 15.6 | N | 23.1 | N | B434D, E449I, F441T, M161P, T387D | N | R470F, E422N, P451I, P585M, D443G | N | Q456S, Q565A |
| 20 | N | N | N | N | N | N | N | N | N | N | N | N | N | N | N | N | N | N |
| 27 | N | N | N | N | 48.1 | 88.6 | 101.3 | 96.3 | 78.1 | 142.4 | N | 72.2 | N | B434D, E449I, F441T, M161P, T387D | N | R470F, E422N, P451I, P585M, D443G | N | Q456S, Q565A |
| 38 | 100 | N | N | N | N | 93.9 | 91.9 | 95.7 | 92.6 | 136.3 | N | 87.3 | N | B434D, E449I, F441T, M161P, T387D | N | R470F, E422N, P451I, P585M, D443G | N | Q456S, Q565A |
| 39 | N | N | 100 | N | 422.5 | 119.2 | 75.0 | 122.1 | 110.7 | 77.5 | N | 173.1 | N | B434D, E449I, F441T, M161P, T387D | N | R470F, E422N, P451I, P585M, D443G | N | A477V, Q456S, Q565A |
| 41 | N | 100 | N | 100 | 78.5 | 79.6 | 104.5 | 98.4 | 96.8 | 102.7 | 54.4 | 92.4 | L21V | B434D, E449I, F441T, M161P, T387D | S80F | R470F, E422N, P451I, P585M, D443G | S84L | Q456S, Q565A |
| 46 | N | N | N | N | 122.0 | 90.9 | 104.9 | 93.9 | 102.2 | 98.3 | N | 107.0 | N | B434D, E449I, F441T, M161P, T387D | N | R470F, E422N, P451I, P585M, D443G | N | Q456S, Q565A |
| 54 | N | N | N | N | N | 182.9 | 89.8 | 57.9 | 77.0 | 45.6 | N | 189.3 | F99Y | B434D, E449I, F441T, M161P, T387D | N | R470F, E422N, P451I, P585M, D443G | N | Q456S, Q565A |
| 78 | N | N | N | N | N | N | N | N | N | N | N | N | N | N | N | N | N | N |
| 79 | N | N | N | N | 900.4 | 90.1 | 102.5 | 96.6 | 99.9 | 114.1 | N | 106.8 | N | B434D, E449I, F441T, M161P, T387D, V90I | N | R470F, E422N, P451I, P585M, D443G | N | Q456S, Q565A |
| 99 | N | N | N | N | 4.9 | 27.2 | 19.3 | 11.7 | 7.8 | 12.7 | 1.0 | 10.8 | N | B434D, E449I, F441T, M161P, T387D | N | R470F, E422N, P451I, P585M, D443G | N | Q456S, Q565A |
| 103 | N | N | N | N | 75.7 | 93.9 | 89.3 | 117.9 | 99.5 | 117.3 | N | 79.9 | N | B434D, E449I, F441T, M161P, T387D | N | R470F, E422N, P451I, P585M, D443G | N | Q456S, Q565A |
| 117 | N | N | N | N | N | N | N | N | N | N | N | N | N | N | N | N | N | N |
| 119 | N | 74.7 | N | N | 45.1 | 101.5 | 94.8 | 95.4 | 98.2 | 117.8 | N | 90.6 | N | B434D, E449I, F441T, M161P, T387D | N | R470F, E422N, P451I, P585M, D443G | N | Q456S, Q565A |
| 124 | N | N | N | N | N | 15.0 | 7.9 | 5.8 | 4.9 | 12.5 | N | 9.4 | N | B434D, E449I, F441T, M161P, T387D | N | R470F, E422N, P451I, P585M, D443G | N | Q456S, Q565A |

*Table S6. Sequence information for antimicrobial resistance determinants. Percent coverage of mobile resistance genes where detected, fold-coverage of chromosomal genes and detected mutations in chromosomal genes. Only genes with mapped sequence data are reported.*
