## Supplementary Table S3 for "Clinical Metagenomic Sequencing for Species Identification and Antimicrobial Resistance Prediction in Orthopaedic Device Infection"

**Table S3. Sample type, culture and histology results and metagenomic sequencing data.** *Comparison of sonication fluid and PPT culture results with species identified after genome coverage filtering by metagenomic sequencing.*

| Sample information | | Culture results | | | | Histology results | Sequencing results | | | | | |
|---|---|---|---|---|---|---|---|---|---|---|---|---|
| Sample number | Orthopedic device type | Sonication species | Sonication CFU/ml count | Tissue culture species | No. of positive tissue samples/total no. samples | Evidence (1) or no evidence (0) of acute infection; results not available (N/A) | Sequencing run batch | Barcode | Total non-human bases | Sequencing species after filtering | Reference genome coverage at 1x (%) | Average reference genome coverage depth (fold) |
| 1 | Knee | *Staphylococcus aureus* | not recorded | *Staphylococcus aureus* | 5/5 | 1 | 1 | 02 | 29025836 | *Staphylococcus aureus* | 94.36 | 8.05 |
| 2 | Orthopedic implant, not otherwise specified | *Staphylococcus aureus* | <5 | *Staphylococcus aureus* | 3/6 | N/A | 1 | 05 | 4728439 | *Staphylococcus aureus* | 51.88 | 1.76 |
| 3 | Metalwork (ulna) | *Enterobacter cloacae* | not recorded | *Enterobacter cloacae* | 4/4 | 1 | 1 | 06 | 23723255 | | | |
| | | | | | | | | | | *Enterobacter ludwigii* | 53.71 | 2.60 |
| | | | | | | | | | | *Cutibacterium acnes* | 59.10 | 2.87 |
| 4 | Knee | negative | | negative | | 0 | 2 | 01 | 1185276 | | | |
| 5 | Metalwork (ankle) | coagulase negative *Staphylococcus* | >49 | coagulase negative *Staphylococcus* | 4/4 | 0 | 2 | 04 | 274201506 | *Staphylococcus caprae* | 89.72 | 51.06 |
| 6 | Knee | *Staphylococcus aureus* | >49 | *Staphylococcus aureus* | 5/5 | 1 | 2 | 08 | 1299546137 | *Staphylococcus aureus* | 93.91 | 337.85 |
| 7 | Metalwork (ankle) | *Streptococcus pyogenes* | >49 | *Streptococcus pyogenes* | 4/4 | N/A | 3 | 05 | 332424257 | *Streptococcus pyogenes* | 93.48 | 161.86 |
| 8 | Knee | *Escherichia coli* | 25-49 | *Escherichia coli* | 6/6 | 1 | 3 | 06 | 2850816 | *Escherichia coli* | 7.04 | 2.76 |
| | | | | *Enterococcus faecium* | 6/6 | | | | | | | |
| 9 | Knee | *Streptococcus dysgalactiae* | >49 | *Streptococcus dysgalactiae* | 5/5 | 1 | 4 | 07 | 136971223 | *Streptococcus dysgalactiae* | 75.52 | 64.64 |
| 10 | Knee (endobutton) | *Staphylococcus aureus* | >49 | *Staphylococcus aureus* | 5/5 | 1 | 4 | 01 | 525272748 | *Staphylococcus aureus* | 97.02 | 151.34 |
| | | *Staphylococcus aureus* | >49 | | | | | | | | | |
| 11 | Knee | negative | | *Staphylococcus epidermidis* | 3/5 | 1 | 5 | 02 | 11965778 | | | |
| 12 | Knee | *Cutibacterium acnes* | >49 | *Cutibacterium acnes* | 3/5 | 1 | 5 | 10 | 1767525541 | *Cutibacterium acnes* | 100.00 | 633.41 |
| 13 | Metalwork (fibula) | *Staphylococcus aureus* | >49 | *Staphylococcus aureus* | 5/5 | 1 | 6 | 01 | 91799578 | *Staphylococcus aureus* | 81.97 | 30.26 |
| 14 | Metalwork (foot) | negative | | negative | | 0 | 6 | 06 | 13920488 | | | |
| 15 | Ankle | negative | | coagulase negative *Staphylococcus* | 2/5 | 0 | 6 | 08 | 31806044 | | | |
| 16 | Hip | *Staphylococcus lugdunensis* | >49 | *Staphylococcus lugdunensis* | 5/5 | 1 | 6 | 09 | 5533753660 | *Staphylococcus lugdunensis* | 98.35 | 1111.58 |
| 17 | Orthopedic implant, not otherwise specified | *Proteus mirabilis* | >49 | *Proteus mirabilis* | 4/5 | 1 | 7 | 01 | 108831184 | | | |
| 18 | Hip | *Staphylococcus epidermidis* | >49 | *Staphylococcus epidermidis* | 4/4 | 1 | 7 | 04 | 2056802094 | *Staphylococcus epidermidis* | 94.87 | 508.97 |
| 19 | Metalwork (radius) | negative | | negative | | 0 | 8 | 05 | 24441840 | | | |
| 20 | Hip | *Staphylococcus aureus* | >49 | *Staphylococcus aureus* | 4/4 | 1 | 8 | 08 | 1114697604 | *Staphylococcus aureus* | 79.95 | 20.65 |
| | | coagulase negative *Staphylococcus* | >49 | coagulase negative *Staphylococcus* | 1/4 | | | | | *Staphylococcus schleiferi* | 91.54 | 28.77 |
| | | | | | | | | | | *Fusobacterium nucleatum* | 79.50 | 363.23 |
| | | | | | | | | | | *Escherichia coli* | 48.24 | 6.83 |
| 21 | Elbow | negative | | negative | | 0 | 9 | 12a | 1257520 | | | |
| 22 | Knee | *Enterococcus faecium* | 25-49 | *Enterococcus faecium* | 6/6 | 1 | 9 | 07 | 4735939669 | *Enterococcus faecium* | 94.00 | 178.33 |
| | | *Staphylococcus epidermidis* | <5 | *Staphylococcus lugdunensis* | 3/6 | | | | | | | |
| | | | | coagulase negative *Staphylococcus* | 4/6 | | | | | | | |
| | | | | *Bacillus circulans* | 1/6 | | | | | | | |
| 23 | Hip | coagulase negative *Staphylococcus* | not recorded | coagulase negative *Staphylococcus* | 5/6 | 1 | 10 | 02 | 206023395 | | | |
| | | | | *Staphylococcus epidermidis* | 2/6 | | | | | | | |
| 24 | Hip | negative | | *Streptococcus dysgalactiae* | 2/3 | 1 | 10 | 10 | 140129518 | | | |
| 25 | Knee | *Enterobacter cloacae* | >49 | *Enterobacter cloacae* | | 1 | 11 | 01 | 5570859830 | *Enterobacter cloacae* | 77.33 | 212.53 |
| | | | | | | | | | | *Enterobacter hormaechei* | 91.86 | 627.67 |
| 26 | Hip | *Staphylococcus epidermidis* | >49 | *Staphylococcus epidermidis* | 2/4 | 1 | 11 | 06 | 5829918 | *Staphylococcus epidermidis* | 6.63 | 16.75 |
| 27 | Elbow | *Staphylococcus aureus* | >49 | *Staphylococcus aureus* | 6/6 | 1 | 13 | 11 | 1940136774 | *Staphylococcus aureus* | 97.00 | 605.71 |
| 28 | Hip | negative | | negative | | 0 | 15 | 10 | 13044263 | | | |
| 31 | Hip | *Pseudomonas aeruginosa* | not recorded | *Pseudomonas aeruginosa* | | 1 | 18 | 09 | 390180 | | | |
| 33 | Knee | *Staphylococcus lugdunensis* | 25-49 | *Staphylococcus lugdunensis* | 6/6 | 1 | 22 | 09 | 2454068563 | *Staphylococcus lugdunensis* | 95.51 | 891.14 |
| 34 | Ankle | *Staphylococcus aureus* | 10-24 | *Staphylococcus aureus* | 5/5 | 1 | 23 | 11 | 8734639 | | | |
| 35 | Metalwork (wrist) | negative | | negative | | 0 | 25 | 02 | 3232588 | | | |
| 36 | Hip | negative | | *Staphylococcus epidermidis* | 1/6 | 0 | 25 | 07 | 1605490 | | | |
| | | | | *Staphylococcus haemolyticus* | 1/6 | | | | | | | |
| 37 | Elbow | negative | | negative | | 0 | 26 | 09 | 903085 | | | |
| 38 | Hip | *Staphylococcus aureus* | >49 | *Staphylococcus aureus* | 5/5 | 1 | 26 | 11 | 3629537539 | *Staphylococcus aureus* | 92.57 | 1237.48 |
| 39 | Knee | *Staphylococcus aureus* | <5 | *Staphylococcus aureus* | 5/5 | 1 | 27 | 02 | 902787550 | *Staphylococcus aureus* | 94.83 | 296.04 |
| 40a | Metalwork (foot) | *Streptococcus dysgalactiae* | not recorded | *Streptococcus dysgalactiae* | 4/5 | 1 | 27 | 10 | 1136875641 | *Streptococcus dysgalactiae* | 88.25 | 455.58 |
| 40b | Metalwork (ankle) | negative | | *Streptococcus dysgalactiae* | 4/5 | 1 | 27 | 11 | 8084158 | *Streptococcus dysgalactiae* | 11.62 | 22.07 |
| 41 | Hip | *Staphylococcus aureus* | >49 | *Staphylococcus aureus* | 5/5 | 1 | 28 | 07 | 994792696 | *Staphylococcus aureus* | 94.47 | 289.40 |
| | | *Staphylococcus aureus* | >49 | *Enterococcus faecium* | 5/5 | | | | | *Enterococcus faecium* | 79.44 | 3.35 |
| 42 | Hip | *Staphylococcus epidermidis* | >49 | *Staphylococcus epidermidis* | 4/4 | 1 | 28 | 09 | 1370087043 | *Staphylococcus epidermidis* | 94.31 | 363.65 |
| 43 | Hip | *Staphylococcus capitis* | >49 | *Staphylococcus capitis* | 5/5 | 1 | 28 | 11 | 229875623 | *Staphylococcus capitis* | 96.56 | 79.82 |
| 44 | Hip | negative | | negative | | 0 | 29 | 05 | 4240858 | *Staphylococcus epidermidis* | 3.23 | 26.25 |
| 45 | Orthopedic implant, not otherwise specified | *Enterobacter cloacae* | 25-49 | *Enterobacter cloacae* | 1/4 | 1 | 29 | 07 | 48466441 | | | |
| 46 | Knee | negative | | *Staphylococcus aureus* | 1/6 | 1 | 29 | 11 | 1687201548 | *Staphylococcus aureus* | 94.60 | 559.43 |
| 47 | Knee | negative | | negative | | 1 | 30 | 02 | 13340874 | | | |
| 48 | Knee | negative | | *Bacillus licheniformis* | 1/4 | 0 | 30 | 09 | 24047 | | | |
| 50 | Cement (wrist) | negative | | negative | | N/A | 32 | 02 | 2797211 | | | |
| 51 | Orthopedic implant, not otherwise specified | negative | | *Staphylococcus capitis* | 4/5 | 1 | 32 | 04 | 68764099 | | | |
| | | | | *Cutibacterium acnes* | 1/5 | | | | | | | |
| 52 | Hip | negative | | negative | | 0 | 32 | 06 | 98383 | | | |
| 53 | Knee | negative | | *Paenibacillus* spp. | 1/5 | 1 | 33 | 06 | 21144292 | *Staphylococcus aureus* | 89.12 | 5.98 |
| | | | | *Peptoniphilus* spp. | 1/5 | | | | | | | |
| 54 | Knee | *Staphylococcus aureus* | <5 | *Staphylococcus aureus* | 5/6 | 1 | 33 | 09 | 780995234 | *Staphylococcus aureus* | 91.44 | 265.63 |
| 55 | Hip | *Fusobacterium* spp. | >49 | *Fusobacterium* spp. | 4/4 | 1 | 34 | 01 | 16386974 | | | |
| 56 | Hip | *Staphylococcus epidermidis* | 25-49 | *Staphylococcus epidermidis* | 4/4 | 1 | 34 | 05 | 158100981 | *Staphylococcus epidermidis* | 95.26 | 47.46 |
| 57 | Knee | *Pseudomonas aeruginosa* | 25-49 | *Pseudomonas aeruginosa* | 5/5 | 1 | 35 | 09 | 288078336 | *Pseudomonas aeruginosa* | 51.03 | 24.18 |
| | | | | *Streptococcus agalactiae* | 1/5 | | | | | | | |
| 58 | Metalwork (spine) | *Cutibacterium acnes* | >49 | *Cutibacterium acnes* | 1/4 | N/A | 36 | 02 | 7208268 | *Cutibacterium acnes* | 55.87 | 3.82 |
| 59 | Hip | *Staphylococcus epidermidis* | 10-24 | *Staphylococcus epidermidis* | 5/5 | 1 | 36 | 05 | 1093265140 | *Staphylococcus epidermidis* | 95.17 | 212.52 |
| | | | | | | | | | | *Corynebacterium segmentosum* | 73.89 | 38.08 |
| 60 | Hip | *Parvimonas micra* | >49 | negative | | 1 | 37 | 02 | 1944541285 | *Parvimonas micra* | 92.06 | 1054.58 |
| 61 | Hip | *Staphylococcus epidermidis* | >49 | *Staphylococcus epidermidis* | 4/6 | 1 | 37 | 11 | 2384177490 | *Staphylococcus epidermidis* | 94.76 | 886.02 |
| 62 | Orthopedic implant, not otherwise specified | *Staphylococcus epidermidis* | 10-24 | *Staphylococcus epidermidis* | 3/4 | 0 | 38 | 02 | 119385954 | | | |
| 63 | Hip | *Streptococcus dysgalactiae* | 10-24 | *Streptococcus dysgalactiae* | 6/6 | 1 | 38 | 05 | 1606983628 | *Streptococcus dysgalactiae* | 92.27 | 632.20 |
| 64 | Metalwork (hip) | negative | | negative | | 0 | 38 | 07 | 72296877 | | | |
| 65 | Metalwork (wrist) | negative | | *Bacillus* spp. | 1/6 | 0 | 39 | 09 | 7229290 | | | |
| 66 | Metalwork (radius) | *Enterobacter* spp. | <5 | *Enterobacter cloacae* | 1/6 | 1 | 39 | 11 | 239403972 | *Enterobacter bugandensis* | 73.88 | 21.56 |
| | | | | | | | | | | *Enterobacter roggenkampii* | 61.20 | 16.81 |
| 70 | Knee | negative | | negative | | 0 | 42 | 02 | 43961733 | | | |
| 71 | Knee | negative | | *Candida albicans* | 1/5 | 1 | 42 | 05 | 16318029 | | | |
| | | | | *Cutibacterium acnes* | 1/5 | | | | | | | |
| 72 | Hip | *Streptococcus dysgalactiae* | 25-49 | *Streptococcus dysgalactiae* | 2/5 | 1 | 42 | 07 | 5366369114 | *Streptococcus dysgalactiae* | 92.37 | 1944.05 |
| 73 | Cement (knee) | negative | | negative | | 0 | 43 | 09 | 85030874 | | | |
| 74 | Hip | *Staphylococcus aureus* | <5 | *Staphylococcus aureus* | 5/6 | 1 | 44 | 02 | 21723140 | | | |
| 75 | Orthopedic implant, not otherwise specified | *Streptococcus agalactiae* | >49 | *Streptococcus agalactiae* | 5/5 | 1 | 44 | 06 | 343724830 | *Streptococcus agalactiae* | 93.98 | 54.07 |
| 76 | Knee | negative | | negative | | 0 | 45 | 10 | 26552307 | | | |
| 77 | Knee | negative | | *Staphylococcus epidermidis* | 5/5 | 1 | 46 | 02 | 13811445 | | | |
| 78 | Orthopedic implant, not otherwise specified | *Staphylococcus aureus* | >49 | *Staphylococcus aureus* | 4/4 | 1 | 46 | 10 | 63833373 | *Staphylococcus aureus* | 53.17 | 13.27 |
| 87 | Hip | *Staphylococcus epidermidis* | 10-24 | *Staphylococcus epidermidis* | 6/7 | 1 | 50 | 06 | 90125872 | *Staphylococcus epidermidis* | 95.80 | 26.38 |
| 88 | Metalwork (ankle) | negative | | negative | | 0 | 50 | 10 | 402165 | | | |
| 89 | Knee | negative | | *Staphylococcus capitis* | 4/5 | 1 | 51 | 02 | 482877 | | | |
| 90 | Knee | *Staphylococcus epidermidis* | <5 | *Staphylococcus epidermidis* | 5/6 | 1 | 51 | 08 | 5350507 | *Staphylococcus epidermidis* | 68.00 | 1.99 |
| 91 | Knee | negative | | negative | | 1 | 51 | 11 | 14520769 | | | |
| 79 | Knee | *Staphylococcus aureus* | >49 | *Staphylococcus aureus* | 6/6 | 0 | 52 | 02 | 3113371295 | *Staphylococcus aureus* | 92.83 | 988.43 |
| | | *Staphylococcus aureus* | >49 | | | | | | | | | |
| 81 | Knee | *Streptococcus oralis* | >49 | *Streptococcus oralis* | 6/6 | 1 | 52 | 06 | 3802687337 | *Streptococcus oralis* | 82.54 | 1375.07 |
| | | | | | | | | | | *Streptococcus* sp. NPS 308 | 67.59 | 121.03 |
| 92 | Metalwork (tibia) | negative | | negative | | 0 | 52 | 09 | 37318038 | | | |
| 95 | Metalwork (tibia) | negative | | negative | | 0 | 53 | 07 | 1982798 | | | |
| 96 | Knee | coagulase negative *Staphylococcus* | <5 | negative | | 0 | 54 | 07 | 329006 | | | |
| 97 | Knee | negative | | negative | | 0 | 55 | 02 | 12377585 | | | |
| 98 | Knee | negative | | *Staphylococcus epidermidis* | 6/6 | 0 | 55 | 05 | 20169729 | | | |
| 99 | Wrist | *Enterobacter cloacae* | not recorded | *Enterobacter cloacae* | 4/4 | 1 | 55 | 10 | 4129774108 | *Enterobacter cloacae* | 75.47 | 15.00 |
| | | *Staphylococcus epidermidis* | not recorded | *Staphylococcus epidermidis* | 4/4 | | | | | *Staphylococcus epidermidis* | 95.02 | 11.82 |
| | | *Dermabacter hominis* | not recorded | *Dermabacter hominis* | 1/4 | | | | | | | |
| | | | | *Staphylococcus aureus* | 1/4 | | | | | *Staphylococcus aureus* | 91.33 | 13.26 |
| | | | | *Aspergillus* spp. | 1/4 | | | | | *Enterobacter hormaechei* | 87.05 | 26.25 |
| | | | | | | | | | | *Anaerococcus mediterraneensis* | 62.46 | 100.79 |
| | | | | | | | | | | *Prevotella intermedia* | 90.86 | 39.83 |
| | | | | | | | | | | *Dermabacter vaginalis* | 58.47 | 2.34 |
| 100 | Knee | *Streptococcus oralis* | >49 | *Streptococcus oralis* | 6/6 | 1 | 56 | 01 | 382506708 | *Streptococcus oralis* | 89.56 | 127.90 |
| 101 | Knee | negative | | negative | | 0 | 56 | 04 | Failed sequencing | | | |
| 103 | Hip | *Staphylococcus aureus* | 10-24 | *Staphylococcus aureus* | 5/5 | 1 | 56 | 08 | 3822239040 | *Staphylococcus aureus* | 94.89 | 1190.12 |
| | | *Staphylococcus aureus* | 25-49 | | | | | | | | | |
| 104 | Hip | *Enterococcus faecalis* | >49 | *Streptococcus oralis* | 5/5 | 1 | 57 | 01 | 10500280 | | | |
| 105 | Hip | *Finegoldia magna* | not recorded | *Finegoldia magna* | 1/6 | 1 | 57 | 04 | Failed sequencing | | | |
| 106 | Elbow | negative | | negative | | N/A | 58 | 01 | 421157 | | | |
| 107 | Knee | *Staphylococcus epidermidis* | 25-49 | *Staphylococcus epidermidis* | 5/5 | 1 | 58 | 04 | 4384221 | *Staphylococcus epidermidis* | 44.77 | 2.45 |
| 108 | Metalwork (femur) | negative | | *Enterococcus faecalis* | 4/4 | 1 | 58 | 11 | 758060 | | | |
| 109 | Hip | *Staphylococcus epidermidis* | <5 | negative | | 1 | 59 | 02 | 205377088 | *Staphylococcus epidermidis* | 93.24 | 51.90 |
| 110 | Hip | negative | | negative | | 0 | 59 | 05 | 17847 | | | |
| 111 | Hip | negative | | negative | | 0 | 59 | 11 | 78396 | | | |
| 112 | Hip | *Escherichia coli* | not recorded | *Escherichia coli* | 3/4 | 1 | 60 | 02 | 1727396384 | *Escherichia coli* | 88.46 | 69.87 |
| | | | | | | | | | | *Cutibacterium acnes* | 97.33 | 383.44 |
| 113 | Metalwork (foot) | negative | | no tissues | | N/A | 60 | 05 | 230081 | | | |
| 114 | Hip | negative | | negative | | 0 | 61 | 02 | 883981 | | | |
| 115 | Knee | *Candida parapsilosis* | 25-49 | | | 0 | 62 | 07 | 176514 | | | |
| | | | | *Staphylococcus aureus* | 1/6 | | | | | | | |
| | | | | coagulase negative *Staphylococcus* | 1/6 | | | | | | | |
| 116 | Hip | *Enterococcus faecium* | >49 | *Enterococcus faecium* | 5/5 | 1 | 62 | 09 | 1693656070 | *Enterococcus faecium* | 92.74 | 362.90 |
| | | *Staphylococcus epidermidis* | <5 | | | | | | | *Staphylococcus epidermidis* | 77.75 | 14.91 |
| 117 | Knee | *Staphylococcus aureus* | 25-49 | *Staphylococcus aureus* | 6/6 | 1 | 62 | 11 | 81141631 | *Staphylococcus aureus* | 43.38 | 36.83 |
| 118 | Ankle | coagulase negative *Staphylococcus* | <5 | negative | | 0 | 63 | 11 | 52967 | | | |
| 119 | Knee | *Staphylococcus aureus* | >49 | *Staphylococcus aureus* | 4/4 | 1 | 63 | 12a | 7350065534 | *Staphylococcus aureus* | 94.46 | 2336.13 |
| 120 | Knee | *Proteus mirabilis* | <5 | *Proteus mirabilis* | 1/4 | 0 | 63 | 07 | 475061540 | | | |
| | | | | *Staphylococcus epidermidis* | 1/4 | | | | | *Staphylococcus epidermidis* | 97.44 | 105.70 |
| | | | | | | | | | | *Corynebacterium segmentosum* | 72.62 | 11.23 |
| 121 | Metalwork (foot) | negative | | *Pseudomonas aeruginosa* | 3/3 | 1 | 64 | 07 | 304828 | | | |
| | | | | coagulase negative *Staphylococcus* | 1/3 | | | | | | | |
| 122 | Knee | *Enterococcus faecalis* | >49 | *Enterococcus faecalis* | 6/6 | 0 | 64 | 08 | 237693 | | | |
| 123 | Shoulder | negative | | negative | | 0 | 64 | 09 | 293931 | | | |
| 124 | Metalwork (ankle) | *Staphylococcus aureus* | 25-49 | *Staphylococcus aureus* | 5/5 | N/A | 64 | 10 | 32223016 | *Staphylococcus aureus* | 94.24 | 10.32 |
| 125 | Elbow | negative | | negative | | 0 | 64 | 11 | 1052995 | | | |
| 102 | Cement (wrist) | negative | | *Stenotrophomonas maltophilia* | 2/2 | N/A | 65 | 07 | 1207867 | | | |
| 126 | Knee | *Pseudomonas aeruginosa* | 10-24 | *Pseudomonas aeruginosa* | 3/3 | 0 | 65 | 09 | 802278 | | | |
| 127 | Metalwork (tibia) | negative | | negative | | 0 | 66 | 01 | 3516271 | | | |
| 128a | Metalwork (ankle) | negative | | *Enterococcus faecalis* | 5/6 | 0 | 66 | 04 | 14932839 | | | |
| | | | | *Klebsiella oxytoca* | 3/6 | | | | | | | |
| | | | | *Finegoldia magna* | 1/6 | | | | | | | |
| 128b | Metalwork (ankle) | negative | | *Enterococcus faecalis* | 5/6 | 0 | 66 | 05 | 37335423 | | | |
| | | | | *Klebsiella oxytoca* | 3/6 | | | | | | | |
| | | | | *Finegoldia magna* | 1/6 | | | | | | | |
